# Therapeutic use of blood products for the treatment of autoimmune hemolytic anemia: A network meta-analysis protocol

**DOI:** 10.1101/2020.01.15.20017657

**Authors:** Jiawen Deng

## Abstract

Autoimmune hemolytic anemia is a rare blood disorder that can result in anemic hypoxia. Currently, red blood cell (RBC) transfusion is the only effective method of treating this condition. We propose a network meta-analysis that investigates whether the use of different types of blood products (e.g. suspended RBC, leukoreduced RBC, washed RBC, etc.) can decrease adverse events, increase the rate of remission and improve lab results, including hemoglobin, RBC, reticulocyte counts, hematocrit and total bilirubin.

## INTRODUCTION

Autoimmune hemolytic anemia (AIHA) is a rare blood disorder characterized by the hemolysis of self red blood cells (RBCs) as a result of the production of autoantibodies[1]. While there are a variety of treatment options available for mediating the effects of AIHA, including corticosteroids, monoclonal antibody rituximab, and surgical splenectomy, patients with AIHA frequently develop anemic hypoxia that cannot be alleviated using these therapies[2,3]. However, because of the presence of autoantibodies, not only are the survival period of transferred RBCs greatly reduced, but transfusion reactions can also occur even in the case of a negative crossmatch[4].

Previous research has shown that the use of leukoreduced or washed erythrocytes may be effective in reducing transfusion reactions by decreasing the amount of antigens in the transfused blood products[5]. We propose to conduct a systematic review and network meta-analysis (NMA) to investigate whether the use of different types of blood products can decrease adverse events, increase the rate of remission and improve lab figures.

## METHODS

We will conduct this network meta-analysis in accordance to the Preferred Reporting Items for Systematic Reviews and Meta-Analyses (PRISMA) incorporating NMA of health care interventions[6]. This study is currently being reviewed for registration on The International Prospective Register of Systematic Reviews (PROSPERO). Any significant amendments to this protocol will be reported and published with the results of the review.

### Eligibility Criteria

#### Types of Participants

We will include adult patients (18 years or older) who have been diagnosed with autoimmune hemolytic anemia, defined as per individual study criteria.

#### Types of Interventions

We will include any whole or part blood product for the analysis. This may include (but not limited to) suspended erythrocytes, leukoreduced erythrocytes or washed erythrocytes. The transfusion of these products may or may not be supplemented by plasma exchange therapy. Concurrent treatments for AIHA such as corticosteroids, rituximab, cyclosporine, etc. are permitted, although their effects will be ignored. If available, we will create an “untreated” treatment arm, consisting of patients who had not received any blood product.

#### Types of Studies

We will include parallel-groups RCTs. If a RCT uses a crossover design, latest data from before the first crossover will be used.

### Primary Outcomes

#### Remission Incidence (n)

We will evaluate incidence of remission based on data collected at the latest follow-up. Definitions of remission will be defined as per individual study criteria. We expect the definitions of remission to be a combination of improvements in clinical symptoms and lab results.

### Secondary Outcomes

#### Lab Results

We will evaluate hemoglobin count (g/L), RBC count (10^12^/L), reticulocyte count (%), hematocrit (%), and total bilirubin (μmol/L) based on the latest lab results.

#### Adverse Events (n)

We will evaluate the incidence of adverse events based on data collected at the latest follow-up. Definitions of adverse events will be defined as per individual study criteria.

### Search Methods for Identification of Studies

#### Electronic Database Search

We will conduct a database search of MEDLINE, EMBASE, Web of Science, CINAHL, and CENTRAL from inception to January 2020. We will use relevant MeSH headings to ensure appropriate inclusion of titles and abstracts (see ***Table 1*** for search strategy).

**Table 1.** MEDLINE Search Strategy

| Database: OVID Medline Epub Ahead of Print, In-Process & Other Non-Indexed Citations, Ovid MEDLINE(R) Daily and Ovid MEDLINE(R) 1946 to Present |  |
| --- | --- |
| Line |  |
| 1 | exp Anemia, Hemolytic, Autoimmune/ |
| 2 | ((Cold or Hot or Warm) adj2 Agglutinin Disease?).ti,ab,kw,kf. |
| 3 | ((Cold or Hot or Warm) adj2 Antibody Disease?).ti,ab,kw,kf. |
| 4 | ((Cold or Hot or Warm) adj2 Antibody H?emolytic Anemia?).ti,a... |
| 5 | Acquired Autoimmune H?emolytic Anemia.ti,ab,kw,kf. |
| 6 | Idiopathic Autoimmune H?emolytic Anemia.ti,ab,kw,kf. |
| 7 | Secondary Autoimmune H?emolytic Anemia.ti,ab,kw,kf. |
| 8 | Autoimmune H?emolytic Anemia/ |
| 9 | AIHA.ti,ab,kw,kf. |
| 10 | WAIHA.ti,ab,kw,kf. |
| 11 | or/1-10 |
| 12 | exp randomized controlled trial/ |
| 13 | exp Randomized Controlled Trials as Topic/ |
| 14 | random*.mp. |
| 15 | Random Allocation/ |
| 16 | ((singl* or doubl* or tripl* or trebl*) adj3 (blind* or mask*)).mp. |
| 17 | double-blind method/ or single-blind method/ |
| 18 | or/12-17 |
| 19 | 11 and 18 |

Major Chinese databases, including Wanfang Data, Wanfang Med Online, CNKI, and CQVIP will also be searched using a custom Chinese search strategy.

#### Other Data Sources

We will also conduct hand search the reference list of previous meta-analyses and NMAs for included articles.

### Data Collection and Analysis

#### Study Selection

We will perform title and abstract screening independently and in duplicate using Rayyan QCRI (https://rayyan.qcri.org). Studies will only be selected for full-text screening if both reviewers deem the study relevant. Full-text screening will also be conducted in duplicate. We will resolve any conflicts via discussion and consensus or by recruiting a third author for arbitration.

#### Data Collection

We will carry out data collection independently and in duplicate using data extraction sheets developed a priori. We will resolve discrepancies by recruiting a third author to review the data.

#### Risk of Bias

We will assess risk of bias (RoB) independently and in duplicate using The Cochrane Collaboration’s tool for assessing risk of bias in randomized trials[7]. Two reviewers will assess biases within each article in seven domains: random sequence generation, allocation concealment, blinding of participants and personnel, blinding of outcome assessment, incomplete outcome data, selective reporting, and other biases (see ***Table 2*** for definitions of RoB domains).

**Table 2.** Definitions of Risk of Bias Domains

| **Risk of Bias Category** | **Definitions** |
| --- | --- |
| Random Sequence Generation | Generation of a random sequence is considered to be adequate if an unpredictable sequence was generated using a random number table or random number generator. It is not adequate to randomize patients using predictable sequences, such as by date of admission or by birth date. |
| Allocation Concealment | Concealment of treatment allocation is considered to be adequate if investigators responsible for patient selection were unable to predict the treatment that the next patient will receive. Adequate allocation concealment methods include sealed, opaque envelopes or centralized randomization. |
| Blinding of Participants and Personnel | Blinding of participants and personnel is considered to be adequate if the investigators report the use of double-blind, triple-blind or quadruple-blind methods. |
| Blinding of Outcome Assessment | Blinding of outcome assessment is considered to be adequate if the investigators assessing the outcome is blinded. This may include blinding technicians, or by recruiting third-party, blinded radiologists to analyze radiographs. |
| Incomplete Outcome Assessment | Handling of incomplete outcome data is considered to be adequate if there is a balanced loss of patients in all treatment arms, or if all patients are included in the analysis (via the intention-to-treat principle). |
| Selective Outcome Reporting | Outcome reporting is considered to be unbiased if the author reported outcomes commonly reported by similar trials, as well as the results of all pre-planned analyses. |
| Other Bias | Other biases that we will evaluate include group similarity at baseline (selection bias), small sample size bias, adequate follow-up time, funding sources, authors' conflicts of interests and the validity of BMD/fracture assessment methods. |

If a majority of domains are considered to be low risk, the study will be assigned a low RoB. Similarly, if a majority of domains are considered to be high risk, the study will be assigned a high RoB. If more than half of the domains have unclear risk or if there are an equivalent number of low and high, low and unclear or high and unclear domains, the study will be assigned an unclear RoB.

### Data Items

#### Bibliometric Data

Authors, year of publication, trial registration, digital object identifier (DOI), publication journal, funding sources and conflict of interest.

#### Methodology

\# of participating centers, study setting, blinding methods, phase of study, enrollment duration, randomization and allocation methods, criteria for remission.

#### Baseline Data

\# randomized, # analyzed, mean age, sex, baseline lab results, follow up duration.

#### Outcomes

\# of patients in remission at the latest follow up, lab results at the latest follow up, # of patients who had experienced at least one adverse event at the latest follow up.

### Statistical Analysis

#### Network Meta-Analysis

We will conduct all statistical analyses using R 3.5.1[8]. We will perform NMAs using the gemtc 0.8-3 library which is based on the Bayesian probability framework[9]. Because we expect significant heterogeneity among studies due to differences in methodology, we will use a random effects model[10].

For remission incidence and adverse event incidence we will report the results of the analyses as risk ratio (RR) with 95% credible intervals (CrIs). For Hb, RBC, reticulocyte counts, hematocrit and total bilirubin, we will report the results as mean differences (MDs) with corresponding 95% CrIs. We will run all network models for a minimum of 100,000 iterations to ensure convergence.

If there are outcomes for which we did not gather enough information to perform an NMA, we will provide a qualitative description of the available data and study outcomes.

#### Treatment Ranking

We will use SUCRA scores to provide an estimate as to the ranking of treatments. SUCRA scores range from 0 to 1, with higher SUCRA scores indicating more efficacious treatment arms[11].

#### Missing Data

We will attempt to contact the authors of the original studies to obtain missing or unpublished data. If we cannot obtain missing standard deviations (SDs), the study will be excluded from the analysis even if the mean was provided.

#### Heterogeneity Assessment

We will assess statistical heterogeneity within each outcome network using I^2^ statistics and the Cochrane Q test[12]. We will consider an I^2^ index ≥ 75% as an indication for serious heterogeneity. If we observe serious heterogeneity, we will explore the sources of heterogeneity using meta-regression analyses.

#### Inconsistency

We will assess inconsistency within the network using the node-splitting method[13].

#### Publication Bias

To assess small-study effects within the networks, we will use a comparison-adjusted funnel plot[14]. We will use Egger’s regression test to check for asymmetry within the funnel plot to identify possible publication bias[15]. The drugs will be sorted according to their efficacy by their SUCRA values, with the assumption that smaller trials tend to favor more efficacious trials.

If we observe significant publication bias, we will perform a sensitivity analysis with limitations on sample sizes.

#### Quality of Evidence

We will use the Confidence in Network Meta-Analysis (CINeMA) web application to evaluate confidence in the findings from our NMA[16]. CINeMA adheres to the GRADE approach for evaluating the quality of evidence by assessing network quality based on six criteria: within-study bias, across-study bias, indirectness, imprecision, heterogeneity and incoherence[17,18]

#### Sensitivity Analyses

We will perform a range of sensitivity/subgroup analyses, with the following limitations:

- Including studies that report the same criteria for evaluating AIHA remission (for the outcome of remission incidence only)
- Including only studies that reported a 24 hour follow up
- Including only studies that reported a 7 day follow up
- Including only studies with a low risk of bias

If we observe significant publication bias, we will perform the following sensitivity analyses:

- Limiting sample size of the included studies to n ≥ 10 in each treatment arm
- Limiting sample size of the included studies to n ≥ 30 in each treatment arm
- Limiting sample size of the included studies to n ≥ 50 in each treatment arm
- Limiting sample size of the included studies to n ≥ 100 in each treatment arm

#### Meta-Regression

If we observe serious heterogeneity in a network, we will perform meta-regression analysis for gender, age, primary/secondary and warm/cold AIHA percentage for that particular network to determine the source of heterogeneity. We will report the results of the meta-regression as a regression coefficient with 95% CrI.

## DISCUSSION

To our knowledge, there are currently no knowledge synthesis regarding the type of blood product to use for treating AIHA. Our proposed study will assist physicians and patients with selecting the best blood product regimen as a concurrent, supplementary therapy to other AIHA treatments.

## Data Availability

All available data are disclosed in the manuscript.

## ACKNOWLEDGEMENTS

None

## AUTHOR STATEMENT

JD made significant contributions to conception and design of the work, drafted the work, and substantially reviewed it.

## REFERENCES

1 Park SH. Diagnosis and treatment of autoimmune hemolytic anemia: classic approach and recent advances. Blood Res 2016; 51: 69–71. doi: 10.5045/br.2016.51.2.69

2 Zanella A, Barcellini W. Treatment of autoimmune hemolytic anemias. Haematologica 2014; 99: 1547–54. doi: 10.3324/haematol.2014.114561

3 Yürek S, Mayer B, Almahallawi M, et al. Precautions surrounding blood transfusion in autoimmune haemolytic anaemias are overestimated. Blood Transfus 2015; 13: 616–21. doi: 10.2450/2015.0326-14

4 Branch DR. Blood Transfusion in Autoimmune Hemolytic Anemias. Lab Med 1984; 15: 402–8. doi: 10.1093/labmed/15.6.402

5 Kim Y, Xia BT, Chang AL, et al. Role of Leukoreduction of Packed Red Blood Cell Units in Trauma Patients: A Review. Int J Hematol Res 2016; 2: 124–9. doi: 10.17554/j.issn.2409-3548.2016.02.31

6 Hutton B, Salanti G, Caldwell DM, et al. The PRISMA extension statement for reporting of systematic reviews incorporating network meta-analyses of health care interventions: checklist and explanations. Ann Intern Med 2015; 162: 777–84. doi: 10.7326/M14-2385

7 Higgins JPT, Altman DG, Gotzsche PC, et al. The Cochrane Collaboration’s tool for assessing risk of bias in randomised trials. BMJ. 2011; 343: d5928–d5928. doi: 10.1136/bmj.d5928

8 Team RC. R: A language and environment for statistical computing. 2015.

9 van Valkenhoef G, Lu G, de Brock B, et al. Automating network meta-analysis. Res Synth Methods 2012; 3: 285–99. doi: 10.1002/jrsm.1054

10 Serghiou S, Goodman SN. Random-Effects Meta-analysis: Summarizing Evidence With Caveats. JAMA 2019; 321: 301–2. doi: 10.1001/jama.2018.19684

11 Mbuagbaw L, Rochwerg B, Jaeschke R, et al. Approaches to interpreting and choosing the best treatments in network meta-analyses. Syst Rev 2017; 6: 79. doi: 10.1186/s13643-017-0473-z

12 Higgins JPT, Thompson SG, Deeks JJ, et al. Measuring inconsistency in meta-analyses. BMJ 2003; 327: 557–60. doi: 10.1136/bmj.327.7414.557

13 van Valkenhoef G, Dias S, Ades AE, et al. Automated generation of node-splitting models for assessment of inconsistency in network meta-analysis. Res Synth Methods 2016; 7: 80–93. doi: 10.1002/jrsm.1167

14 Chaimani A, Higgins JPT, Mavridis D, et al. Graphical tools for network meta-analysis in STATA. PLoS One 2013; 8: e76654. doi: 10.1371/journal.pone.0076654

15 Peters JL, Sutton AJ, Jones DR, et al. Comparison of two methods to detect publication bias in meta-analysis. JAMA 2006; 295: 676–80. doi: 10.1001/jama.295.6.676

16 Institute of Social and Preventive Medicine, University of Bern. CINeMA: Confidence in Network Meta-Analysis. 2017.

17 Salanti G, Del Giovane C, Chaimani A, et al. Evaluating the quality of evidence from a network meta-analysis. PLoS One 2014; 9: e99682. doi: 10.1371/journal.pone.0099682

18 Nikolakopoulou A, Higgins JPT, Papakonstantinou T, et al. Assessing Confidence in the Results of Network Meta-Analysis (Cinema). doi: 10.1101/597047

19 Abstracts of the Global Evidence Summit. In: Abstracts of the Global Evidence Summit. Wiley 2017.doi: 10.1002/14651858.CD201702

